# Results of the basic-VRS trial: clinical outcomes and cost-effectiveness of basic low vision rehabilitation in Portugal

**DOI:** 10.1101/2022.07.29.22278192

**Authors:** Laura Hernández-Moreno, Hugo Senra, Ana Patricia Marques, Natacha Moreno Perdomo, Antonio Filipe Macedo

## Abstract

**Purpose:** The aim of this study was twofold: i) to investigate and characterize the clinical impact of vision rehabilitation in patients with vision impairment, and ii) to investigate the cost-effectiveness of a basic vision rehabilitation service in Portugal.

**Methods:** Patients diagnosed with age-related macular degeneration or diabetic retinopathy and visual acuity in the range 0.4 to 1.0 logMAR in the better-seeing eye were recruited. Participants were randomised to one of the study arms consisting of immediate intervention and delayed intervention. The intervention included: new refractive correction, optical reading aids, in-office training and advice about modifications at home. Visual ability, health-related quality-of-life and costs of the intervention were measured. Economic analysis was performed to evaluated if the intervention was cost-effective. The trial compared the outcomes 12-weeks after the start in both arms.

**Results:** Of the 46 participants, 34(74%) were diagnosed with diabetic retinopathy, 25(54%) were female and the mean age was 70.08 yrs (SD=8.74). In the immediate intervention arm visual ability was 0.28 logits (SD=1.14) at baseline and it increased to 0.91logits (SD=1.24) after the intervention (p<0.001). Changes in the delayed intervention arm were not statistically significant (mean improvement = 0.10 logits, SE=0.11, p=0.95). Acuity in the better seeing eye, near acuity and critical print size also improved during the study. The mean cost of the intervention was €118.79 (SD=24.37). Incremental cost-effectiveness ratio using the EQ-5D-5L index value was €3322/QALY and €1235/QALY when using near acuity.

**Conclusions:** The current study gives evidence of positive clinical impact of a basic vision rehabilitation intervention and that a basic vision rehabilitation service is cost-effective. These findings are important to clinical and rehabilitation practices and for planning vision rehabilitation services.

## Introduction

Globally millions of people live with some level of visual impairment (VI) [1-3]. VI is a common cause of disability affecting patients’ well-being, mental health, activities of daily living and social functioning (e.g. independence or difficulties to find job) [4-10]. VI leads to significant economic burdens due to direct costs (inpatient and outpatient care) and indirect cost (informal care or productivity losses) [7, 8, 11-14]. For example, reduced independence to perform activities of daily living can lead to less job opportunities due to reduced ability to work [15-17]. Vision rehabilitation can be effective in tackling these limitations and promoting independent living and autonomy in people with VI [18].

Vision rehabilitation (VR) can be defined as a mixture of health, educational and social interventions, whose ultimate goal is to reduce the negative impact of VI. The aim of these services is to improve visual ability (the ability to perform tasks that rely on vision) [19, 20] and other aspects associated with VI such as the psychosocial burden [18, 21]. VR works by enhancing visual function which includes, for example, the use of assistive devices or changes in the visual environment such as improved lighting. VR often requires the acquisition of new skills such as handling assistive devices (e.g. haptic devices such as BrainPort [22]) or accessibility features in ordinary electronic devices [23]. With rehabilitation, activities that rely on vision are expected to become easier to perform even without improvement in visual acuity [6, 21, 24, 25]. Previous studies suggested that, in general, patients are satisfied with VR and acknowledge its benefits for quality of life and functioning [5, 26].

Lately our group has carried out some studies showing that VI is common in Portugal and that many people are still struggling to cope with the condition [7-9, 27-30]. In general, there is still lack of studies examining the actual benefits of VR for people with VI. This lack of evidence entails barriers to the development of vision rehabilitation systems (VRS) [31-34]. Magnifiers, for example, have been dispensed in many hospitals regularly since, at least, 1970 [35]. However, a systematic review showed that better evidence on the benefits of using magnifiers is necessary [26]. In addition, despite the widespread prescription of magnifiers, there is still insufficient evidence on the effect of different types of low vision aids on reading performance [36, 37]. In short, more research addressing the actual impact of VR on patients’ functioning is needed to inform clinicians and rehabilitation professionals on the best practice for visually impaired patients.

The aim of this trial was (1) to assess the effect of a basic vision rehabilitation intervention in visual ability in people with impaired vision, (2) to report the cost-effectiveness of a basic vision rehabilitation service provided in a Portuguese setting. In this study we tested the hypothesis that a basic vision rehabilitation intervention improves patients’ ability to perform activities of daily living and it is cost-effective [38].

## Methods

This study is part of a clinical trial addressing the cost-effectiveness of a basic vision rehabilitation service in Portugal (registration number: ISRCTN10894889). The study was approved by the Ethics Committee for Life Sciences and Health of the University of Minho (SECVS 147/2016), and by the Hospital Santa Maria Maior’s ethics committee. Portuguese data protection authority approval number: 7012/ 2017.

Patients attending outpatient appointments at the department of ophthalmology at Hospital Santa Maria Maior E.P.E (Barcelos, Portugal) were invited to participate in this study. The inclusion criteria were: i) visual acuity between 0.4 and 1.0 logMAR in the better-seeing eye; ii) primary diagnosis and main cause of vision loss should be diabetic retinopathy or age-related macular degeneration; iii) 18 years or older and iv) living in the community (not living in any type of institution). The exclusion criteria were: i) cognitive impairment based on scores on mini-mental state examination, ii) communication problems due to, for example, hearing impairment, or inability to speak Portuguese; iii) inability to read due to low level of education; iv) inability to attend the requested appointments at the study setting. For those accepting to take part, demographic and clinical information was collected. Cognitive status was assessed using the Portuguese version of The Mini-Mental State Examination (MMSE) [39].

The study design adopted was a parallel group randomized controlled trial. Participants were allocated to an immediate intervention group or arm (IMI), or to a delayed intervention group or arm (DEI). The IMI group received the intervention in the first visit (baseline), the DEI group was used as to control for a possible effect of ‘attention to the problem’ and participants were put in a waiting list – participants in this group only received the intervention in the second visit - 12 weeks after the first visit.

The basic vision rehabilitation intervention (VRI) consisted of 3 main components: (1) prescription, when necessary, of the best refractive correction for distance vision (2) prescription of magnification for reading (near glasses or handheld magnifiers) and (3) instructions and training. The detailed procedure for each step of the intervention has been published as part of the study protocol – for more details readers are referred to our previous publication [38].

### Main outcome measure and vision measures

#### Main outcome

Visual ability was measured with the Portuguese version of Massof Activity Inventory (AI) [7, 8, 40, 41]. The AI consists of a hierarchal structure in which specific cognitive and motor visual tasks (e.g., pouring or mixing without spilling) underlie more global goals (e.g., preparing meals) [42, 43]. Disabilities occur when an individual reports difficulty in achieving important goals. Goals are split between three objectives: social functioning, recreation and daily living and associated with four classes of function: reading, visual motor (also called manipulation), visual information (also called seeing), and mobility [42, 43]. Difficulties achieving a goal are said to depend on the difficulty experienced in the tasks that underlie each goal. Respondents first rate the importance of each goal with four possible responses ranging from “not important” to “very important”. Goals rated “not important” are skipped, and as such are not considered in the final visual ability score as these are not relevant to the person’s daily life. For goals rated “slightly important” or above, participants are asked to rate difficulty on a five-point scale ranging from “not difficult” to “impossible to do” [42, 43].

#### Vision outcomes

Distance VA was measured with ETDRS charts, in a dim light room using an internally illuminated cabinet (model 2425E, (Precision Vision, IL, USA)). Near VA was assessed with near version of the ETDRS charts (https://www.precision-vision.com/). VA was measured monocularly at distance and binocularly at near, at both distances a letter-by-letter scoring was used [41, 44]. Testing distance was adjusted according to the severity of vision loss, final acuity scores reported were adjusted to standard distances (4 meters at distance and 40 cm at near).

Reading was tested to determine vision-related reading difficulties. Specifically, we measured: (1) reading acuity (RA), (2) maximum reading speed (MRS), and (3) critical print size (CPS) using the Portuguese version of Minnesota Low-Vision Reading Test (MNread test) [45-47]. Reading was measured binocularly at 40cm or 20cm according to the needs. After the intervention, reading was assessed with the prescribed aid.

Contrast sensitivity was assessed binocularly at 40cm with near correction using the MARS test (https://www.marsperceptrix.com/) which has a gradual letter-by-letter contrast. Illuminance on the surface of the test was approximately 330 lux. Participants were encouraged to respond until two consecutive letters were read incorrectly, scoring was performed according to the test instructions.

### Economic evaluation

#### Measures of costs

Rehabilitation costs were used for the cost estimation. This estimation includes the hospital costs, distance glasses (when necessary) and near glasses or handheld magnifiers. The hospital costs included overheads for facilities and equipment and optometrist’s time. For the cost of distance glasses, near glasses and handheld magnifiers we used as guidelines the price recommended for the public for each case [38].

#### Measures of effectiveness

Health-related quality of life (HRQOL) was accessed with the EuroQol 5 dimensions, 5-point response scale questionnaire (EQ-5D-5L). This questionnaire comprises five dimensions which have five possible levels of response. Three dimensions are related to function (Mobility, Self-Care and Usual Activities) and the other two describe feelings (Pain/Discomfort and Anxiety/Depression) [48, 49]. Utility index values used here were obtained using index value set calculators obtained from https://euroqol.org/ that use valuations of health states in England.

Near VA values were converted to quality-adjusted life year (QALYs) through ophthalmic utility values. As suggested by others in this procedure participant’s near VA is converted to utilities [50, 51], the reference for near vision ophthalmic utility value was the value of time trade-off utility values for patients with ocular diseases [52].

#### Economic and sensitivity analysis

Economic analysis was conducted from the healthcare perspective. All costs given in euros for year 2020. EQ-5D-5L index and near vision ophthalmic utility value were measured before the intervention and 12 weeks after to capture the effect of the intervention. To know if the intervention was cost-effective the incremental cost-effectiveness ratio (ICER) was computed using the expression:

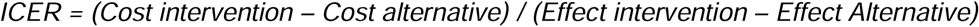

From the IMI group we obtained the “cost intervention” which are the rehabilitation costs and the “effect intervention” which are the utility values from EQ-5D-5L and from the near vision ophthalmic utility value after 12 weeks after the intervention. From the DEI group we obtained the “cost alternative” which was zero (because the group was in a waiting list) and the “effect alternative” or “placebo”.

To assess whether the intervention was cost–effective, the threshold used was based on the Portuguese per-capita Gross Domestic Product (GDP) of €19,431 for year 2020 [53]. However, the World Health Organization’s Commission on Macroeconomics in Health suggested that cost–effectiveness thresholds should be three times the per-capita GDP [54] which gives a cost– effectiveness threshold of €58,293. To determine confidence intervals to our cost-effectiveness we used a procedure used in previous studies [55, 56]. In short, we used bootstrapping with 5 000 replications for the costs and for the effects in both groups to generate 95% confidence intervals around the ICER estimates. Cost-effectiveness planes were plotted to show the distribution of costs and effects. Cost-effectiveness acceptability curves to show the probability of cost-effectiveness at a range of thresholds were also plotted.

Adjusted intervention cost sensitivity analyses were performed to evaluate uncertainty. For that we computed the ICER for our sample and then computed the ICER after altering the costs of the intervention. In one analysis we used the highest cost for the equipment (€220.70) in all participants and in other the lowest cost (€103.12). The use of the mean for ICER calculation is almost unanimous [57, 58], but using the median has been suggested since it is less sensitive to outliers or erroneous data – which happens often in monetary data [57]. In the current study we used both, mean and median.

### Statistical analysis

Descriptive statistics and hypothesis testing were performed according to the type of variable (continuous or discrete) and its distribution (normal or skewed). Kolmogorov-Smirnov test was performed to assess normality. Rasch analysis was used to analyse results of the AI, the analysis was conducted using the Andrich rate scale model [59] for polytomous data with Winsteps software (v. 4.4.0) to compute person measures of visual ability [60]. The effect of time and group on trial outcomes was tested using linear mixed models (LMM) using PROCMIX in SAS software (R: 3.8, SAS Institute Inc., NC, USA). For the main outcome measure, visual ability was normalized by subtracting the AI person measure at week 1 from all measures, that corresponds to a baseline visual ability of 0 for all participants. For this model visual ability was defined as “dependent variable”, participants were defined as “random factors” or “group specific effects”. Explanatory factors or “fixed factors” were: “group” (IMI = immediate intervention or DEI=delayed intervention) and time (1, 12 and 36 for the IMI group and 1, 12 and 24 for the DEI group). Similar models were performed for other trial outcomes, only the dependent variable was changed. Statistical significance was set at p value less than 0.05.

## Results

According to the initial protocol, the estimated sample size to detect a significant difference in visual ability measured with the activity inventory at 12 weeks was 22 per arm [38]. A total of 59 patients were invited to participate, from these 46 accepted to take part in the study. Socio-demographic and clinical characteristics of the 46 participants that started in the study are summarized in Table 1.

**Table 1.** Socio-demographic characteristics of the participants at baseline, dropouts included.

| Variable | Group |  |
| --- | --- | --- |
|  | IMI | DEI |
| **Total n** | 23 | 23 |
| **Age**, mean (SD), in years | 72.61 (13.00) | 70.08 (8.74) |
| **Comorbidities**, mode (Range) | 2 (5) | 4 (6) |
| **Years with VI**, mean (SD), in years | 2.72 (1.85) | 3.64 (3.79) |
| **Presenting VA better eye**, mean (SD), in logMAR | 0.71 (0.20) | 0.66(0.30) |
| **Sex** |  |  |
| • Female, n (%) | 10 (43%) | 15 (63%) |
| • Male, n (%) | 13 (57%) | 9 (37%) |
| **Diagnosis (primary cause of VI)** |  |  |
| • diabetic retinopathy (DR), n (%) | 16 (70%) | 18 (78%) |
| • age-related macular degeneration (AMD), n (%) | 7 (30%) | 5 (22%) |
| **Living** |  |  |
| • With others, n (%) | 19 (82.6%) | 21 (91%) |
| • Alone, n (%) | 4 (17.4%) | 2 (9%) |
| **Education** |  |  |
| • 4 or less years, n (%) | 9 (39%) | 9 (39%) |
| • 6 to 9 years, n (%) | 8 (35%) | 11 (48%) |
| • 12 years n (%) | 4 (17%) | 1 (4%) |
| • University or more, n (%) | 2 (9%) | 2 (9%) |

*IMI - immediate intervention group, DEI - delayed intervention group, VA – visual acuity, VI- visual impairment, SD – standard deviation*

During the duration of the study, which was mostly conducted during the COVID-19 pandemic, there were 10 dropouts (21%) related to: inadaptation to the low vision aids, vision improved due to medical treatments or participants failed to show at follow-up assessments. We considered dropout when participants failed to complete the first follow-up after the intervention (week 12 in the IMI arm or week 24 in the DEI arm).

The time with the optometrist during the vision rehabilitation intervention was approximately 90 minutes (questionnaire administration excluded). The reading aids prescribed compromised a total of 17 new pair of glasses (3 for distance and 14 for near) and 23 LED-illuminate handheld magnifiers. The mean power of the reading aids was 10.0D (SD= 5.0), the median for near glasses was 6D and 12D for the handheld magnifiers.

### Main outcome measure of the trial - visual ability

The mean visual ability in the IMI (n=23) before the intervention at baseline or week 1 was 0.28 logits (SD=1.14) and it increased (n=21 at 12 weeks) to 0.91 logits (SD=1.24) after the intervention. In the DEI group (n=23) the mean visual ability at baseline (week 1) was 0.71 logits (SD=1.30) and it changed (n=16 at 12 weeks) to 0.45 logits (SD=8.88) after the 12-weeks waiting period. Visual ability values were normalized before statistical analysis. A linear mixed model (LMM) with visual ability as dependent variable revealed a main effect of time, given in weeks, (F(3, 69) = 41.16, p<0.001) and an interaction time×group (F(1,69)=6.54, p=0.012), The effect of group was not statistically significant (p=.059). Comparisons within and between groups are summarized in Table 2, bold-font rows correspond to the main results of the trial after 12 weeks whose values are shown in Figure 1.

**Table 2.** Summary of the pairwise comparisons for the interaction time×group and the AI changes within group over time. The comparisons indicate that the differences between week 1 and week 12 for DEI (comparison A) were not statistically significant whilst the differences were statistically significant for the IMI after the intervention (comparison B) and the groups showed statistically significant differences (comparison C).

| Group | Week | Group | Week | MD | SE | DF | t Value | Pr > \|t\| | Adj | Adj P |
| --- | --- | --- | --- | --- | --- | --- | --- | --- | --- | --- |
| **DEI** | **1** | **DEI** | **12** | **-0.1024** | **0.1199** | **69** | **-0.85** | **0.3963<sup>A</sup>** | **T-K** | **0.9560** |
| DEI | 1 | DEI | 24 | -0.6480 | 0.07298 | 69 | -8.88 | <.0001 | T-K | <.0001 |
| **DEI** | **1** | **IMI** | **1** | **0** | **0.1130** | **69** | **-0.00** | **1.0000** | **T-K** | **1.0000** |
| DEI | 1 | IMI | 12 | -0.5233 | 0.1157 | 69 | -4.52 | <.0001 | T-K | 0.0003 |
| DEI | 1 | IMI | 36 | -0.6872 | 0.1170 | 69 | -5.88 | <.0001 | T-K | <.0001 |
| DEI | 12 | DEI | 24 | -0.5457 | 0.1255 | 69 | -4.35 | <.0001 | T-K | 0.0006 |
| DEI | 12 | IMI | 1 | 0.1024 | 0.1225 | 69 | 0.84 | 0.4065 | T-K | 0.9599 |
| **DEI** | **12** | **IMI** | **12** | **-0.4210** | **0.1250** | **69** | **-3.37** | **0.0012<sup>C</sup>** | **T-K** | **0.0151** |
| DEI | 12 | IMI | 36 | -0.5848 | 0.1262 | 69 | -4.63 | <.0001 | T-K | 0.0002 |
| DEI | 24 | IMI | 1 | 0.6480 | 0.1192 | 69 | 5.44 | <.0001 | T-K | <.0001 |
| DEI | 24 | IMI | 12 | 0.1247 | 0.1217 | 69 | 1.02 | 0.3092 | T-K | 0.9082 |
| DEI | 24 | IMI | 36 | -0.03915 | 0.1230 | 69 | -0.32 | 0.7512 | T-K | 0.9995 |
| **IMI** | **1** | **IMI** | **12** | **-0.5233** | **0.1128** | **69** | **-4.64** | **<.0001<sup>B</sup>** | **T-K** | **0.0002** |
| IMI | 1 | IMI | 36 | -0.6872 | 0.1020 | 69 | -6.74 | <.0001 | T-K | <.0001 |
| IMI | 12 | IMI | 36 | -0.1638 | 0.06762 | 69 | -2.42 | 0.0180 | T-K | 0.1628 |

MD= mean difference for the AI between the pair Group-Week in the first 2 columns and the pair Group-Week in column 3 and 4; SE = standard error for the MD; T-K= Tukey-Kramer procedure; IMI - immediate intervention group, DEI - delayed intervention group.

**Figure 1:**
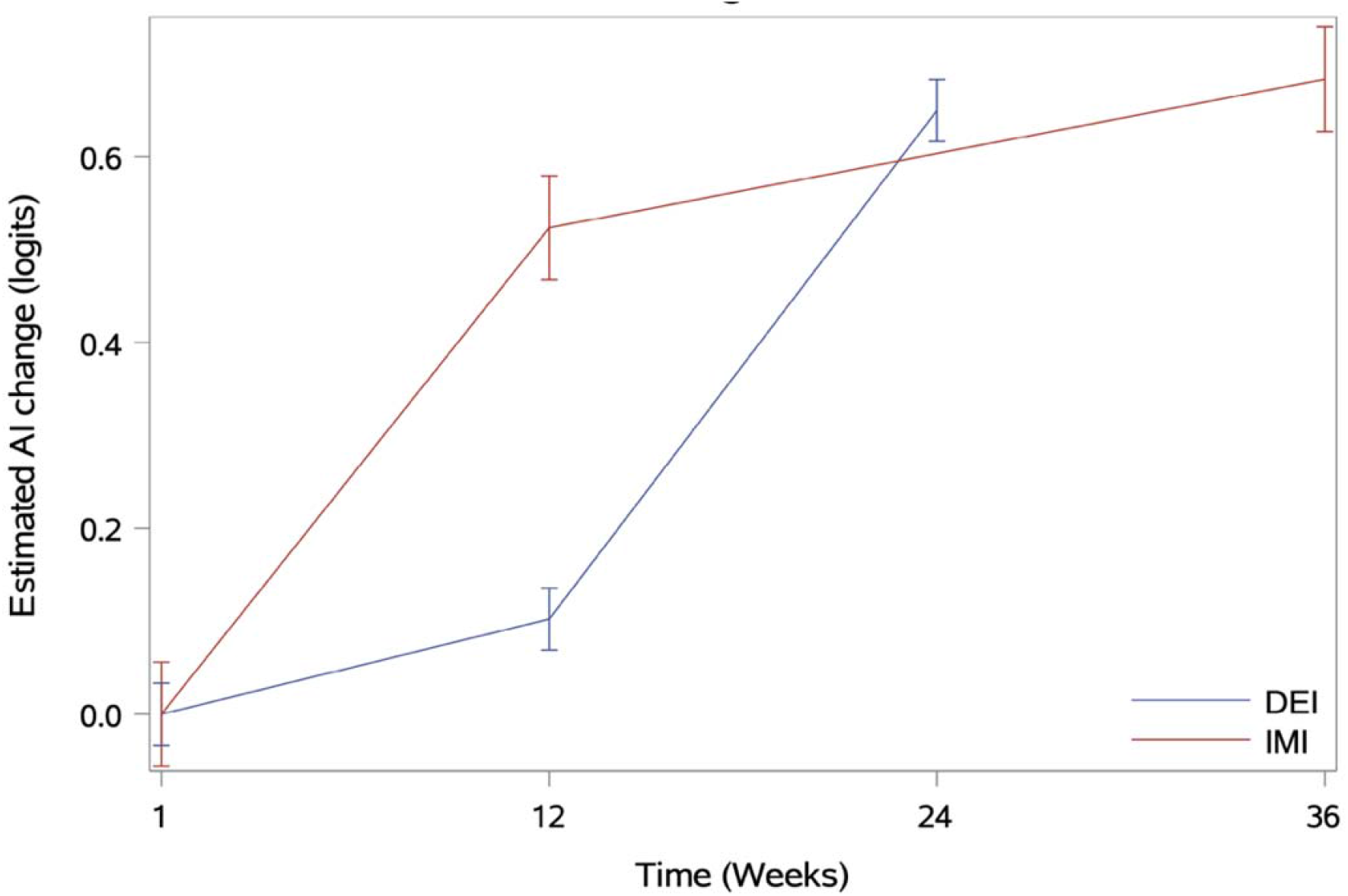
Estimated mean AI change and standard errors for the change for both groups over time. IMI - immediate intervention group, DEI - delayed intervention group. This cross-over design uses the DEI group as control group - between weeks 1 and 12, DEI group was in a waiting list for a low vision intervention but did not receive any attention. The IMI group received a low vision rehabilitation intervention at week 1 and AI changes were assessed at 12 weeks - that assessment corresponds to the main outcome of the trial. Between weeks 12 and 36 weeks the IMI group did not receive any attention and this assessment at 36 weeks was performed to investigate if the benefits of the rehabilitation persisted.

### Clinical changes with rehabilitation

Descriptive statistics for visual outcomes are given in Table 3. A LMM with VA in the better seeing eye as dependent variable revealed a statistically significant effect for factor time (F(3,69)=3.63, p<0.017), the effects of interaction time×group (p<0.54) and factor group (p=0.07) were not statistically significant. These results show that distance acuity in the better seeing eye improved with time for both groups.

**Table 3.**
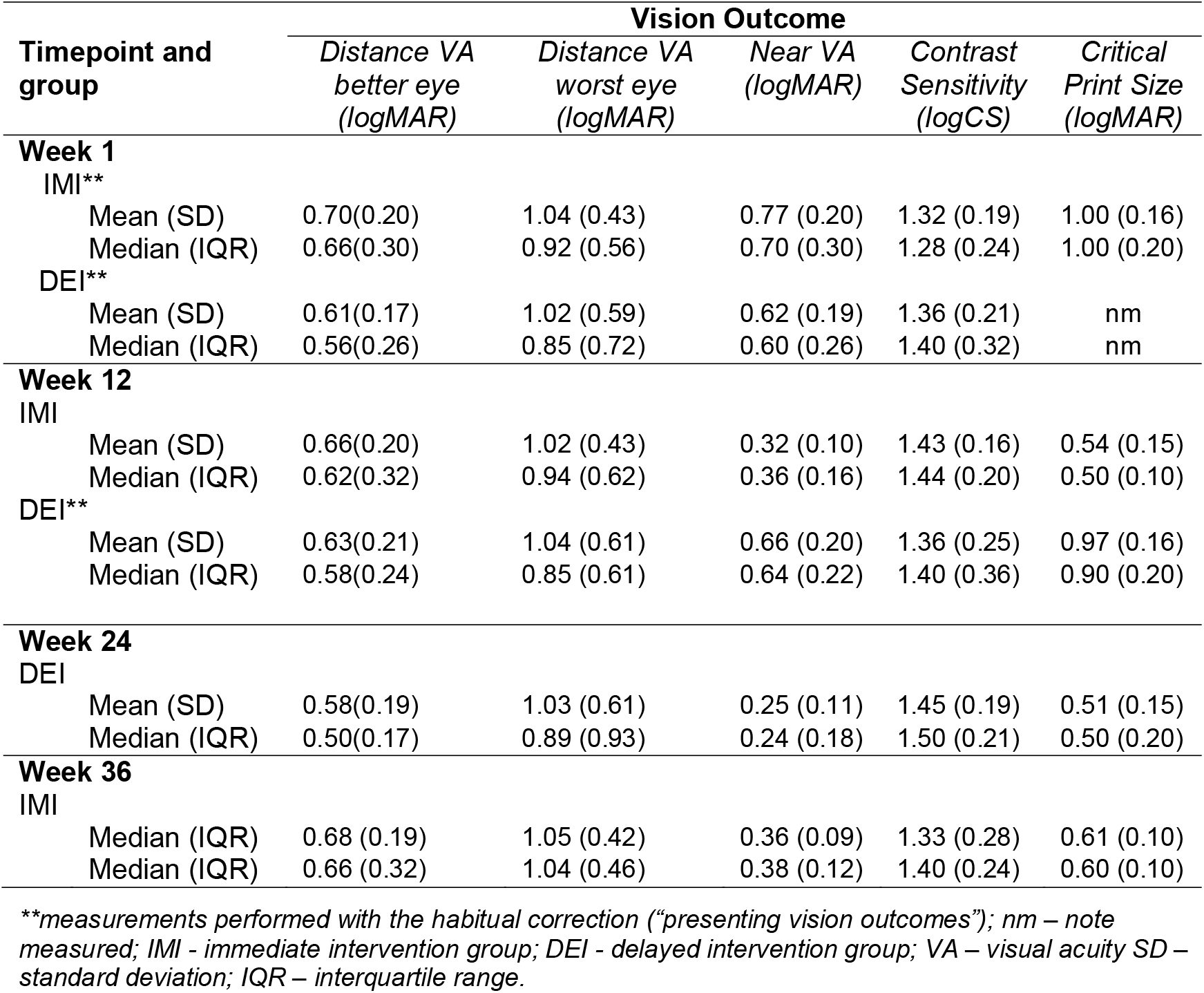
Vision outcomes for IMI and DEI at different timepoints.

| Timepoint and group | Vision Outcome |  |  |  |  |
| --- | --- | --- | --- | --- | --- |
|  | Distance VA better eye (logMAR) | Distance VA worst eye (logMAR) | Near VA (logMAR) | Contrast Sensitivity (logCS) | Critical Print Size (logMAR) |
| **Week 1** |  |  |  |  |  |
| IMI** |  |  |  |  |  |
| Mean (SD) | 0.70(0.20) | 1.04 (0.43) | 0.77 (0.20) | 1.32 (0.19) | 1.00 (0.16) |
| Median (IQR) | 0.66(0.30) | 0.92 (0.56) | 0.70 (0.30) | 1.28 (0.24) | 1.00 (0.20) |
| DEI** |  |  |  |  |  |
| Mean (SD) | 0.61(0.17) | 1.02 (0.59) | 0.62 (0.19) | 1.36 (0.21) | nm |
| Median (IQR) | 0.56(0.26) | 0.85 (0.72) | 0.60 (0.26) | 1.40 (0.32) | nm |
| **Week 12** |  |  |  |  |  |
| IMI |  |  |  |  |  |
| Mean (SD) | 0.66(0.20) | 1.02 (0.43) | 0.32 (0.10) | 1.43 (0.16) | 0.54 (0.15) |
| Median (IQR) | 0.62(0.32) | 0.94 (0.62) | 0.36 (0.16) | 1.44 (0.20) | 0.50 (0.10) |
| DEI** |  |  |  |  |  |
| Mean (SD) | 0.63(0.21) | 1.04 (0.61) | 0.66 (0.20) | 1.36 (0.25) | 0.97 (0.16) |
| Median (IQR) | 0.58(0.24) | 0.85 (0.61) | 0.64 (0.22) | 1.40 (0.36) | 0.90 (0.20) |
| **Week 24** |  |  |  |  |  |
| DEI |  |  |  |  |  |
| Mean (SD) | 0.58(0.19) | 1.03 (0.61) | 0.25 (0.11) | 1.45 (0.19) | 0.51 (0.15) |
| Median (IQR) | 0.50(0.17) | 0.89 (0.93) | 0.24 (0.18) | 1.50 (0.21) | 0.50 (0.20) |
| **Week 36** |  |  |  |  |  |
| IMI |  |  |  |  |  |
| Median (IQR) | 0.68 (0.19) | 1.05 (0.42) | 0.36 (0.09) | 1.33 (0.28) | 0.61 (0.10) |
| Median (IQR) | 0.66 (0.32) | 1.04 (0.46) | 0.38 (0.12) | 1.40 (0.24) | 0.60 (0.10) |

\*\*measurements performed with the habitual correction ("presenting vision outcomes"); nm – note measured; IMI - immediate intervention group; DEI - delayed intervention group; VA – visual acuity SD – standard deviation; IQR – interquartile range.

LMM for near VA as dependent variable revealed statistically significant effect of main factor time (F(3,69)=49.77, p<0.001) and for interaction time×group (F(1,69)=82.61, p<0.001), the effect of group was not statistically significant (p=0.45). These results show that near acuity improved with time for both arms but the interaction indicates that the changes were due to the intervention that was delivered at different times to each arm.

For contrast sensitivity as dependent variable none of the effects tested with LMM was statistically significant. For critical print size as dependent variable the main effect time in weeks was statistically significant (F(3,53)=77.82, p<0.001). These findings reveal that the intervention failed to improve contrast sensitivity but succeed at improving critical print size.

### Economic evaluation

#### Effectiveness and costs of the intervention

The intervention was effective at providing additional QALYs. Using the EQ-5D-5L index value, for the IMI group the median QALY gain of 0.102 (IQR=0.169) after 12 weeks was statistically significant (Wilcoxon-test, Z=-2.670, p=0.008). The median QALY change of 0.00 (IQR=0.287) for the DEI group during the 12 weeks waiting was not statistically significant (p=0.477). The difference in effect between groups (effect = (EQ-5D-5L index value at 12 weeks) <u>minus</u> (EQ-5D-5L index value at week 1) for each participant) at 12 weeks was statistically significant (Mann–Whitney U test, Z=-2.007, p=0.045).

Using near acuity ophthalmic utility value, for the IMI group the mean improvement in QALY of 0.077 (SD=0.072) after 12 weeks was statistically significant (paired t-test t(20)=-5.217, p<0.001.) The mean reduction in QALY of -0.008 (SD=0.041) after 12 weeks for the DEI was not statistically significant (p=0.490). The difference in effect between groups at 12 weeks was statistically significant (independent t-test, t(36)= 4.611, p<0.001).

The mean cost of the intervention was €118.79 (SD=24.37), range from €103.12 to €199.27. The costs of the intervention included €31.23 for hospital costs for each participant, the mean cost for distances glasses was €84 and the mean cost for near aids (near glasses or LED-illuminate handheld magnifiers) was €80.71 (SD=7.87).

#### ICER - Cost-effectiveness results

Table 4 summarizes the effect and ICER results for the 5000 bootstrap replications. The ICER obtained through the EQ-5D-5L index value was €3322.46/QALY and the one obtained through the near acuity ophthalmic utility value was €1235.40/QALY. Based on Portuguese per-capita GDP of €19431 the intervention can be considered cost-effective.

**Table 4.**
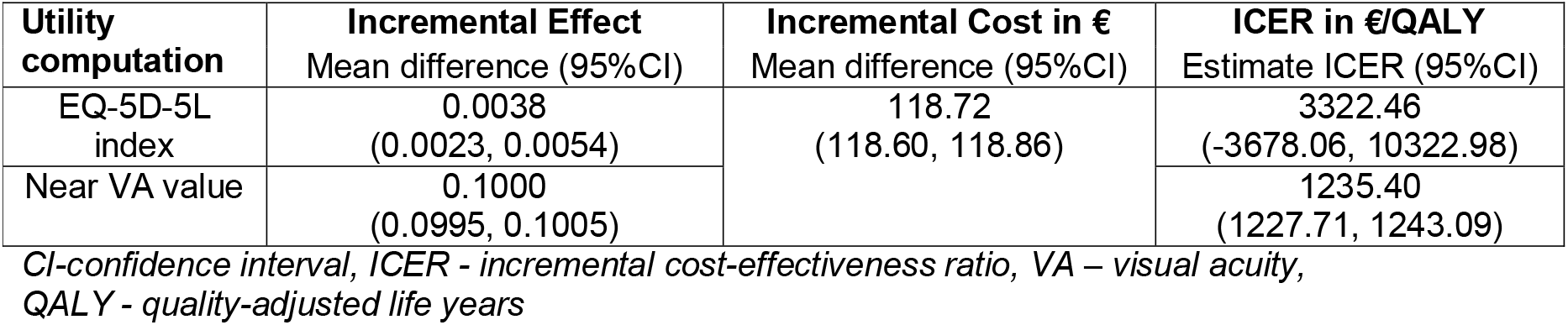
Economic analysis: incremental effect and cost and ICER estimation from bootstrapping.

| Utility computation | Incremental Effect<br>Mean difference (95%CI) | Incremental Cost in €<br>Mean difference (95%CI) | ICER in €/QALY<br>Estimate ICER (95%CI) |
| --- | --- | --- | --- |
| EQ-5D-5L index | 0.0038<br>(0.0023, 0.0054) | 118.72<br>(118.60, 118.86) | 3322.46<br>(-3678.06, 10322.98) |
| Near VA value | 0.1000<br>(0.0995, 0.1005) |  | 1235.40<br>(1227.71, 1243.09) |

*CI*-confidence interval, *ICER* - incremental cost-effectiveness ratio, *VA* – visual acuity, *QALY* - quality-adjusted life years

**Table 5.** Sensitivity analysis adjusted by cost

| Basis for utility computation |  | Real cost ICER in €/QALY | Lowest cost ICER in €/QALY | Highest cost ICER in €/QALY |
| --- | --- | --- | --- | --- |
| EQ-5D-5L index value | Median | 1118.82 | 8501.24 | 2174.38 |
|  | Mean | 9977.54 | 1015.96 | 18194.56 |
| Near vision ophthalmic utility value | Median | 1336.00 | 1213.18 | 2596.47 |
|  | Mean | 1434.32 | 1222.09 | 2615.55 |

*QALY – quality-adjusted life years, ICER - Incremental cost-effectiveness ratio, VA – visual acuity.*

Figure 2 shows the cost-effectiveness planes and the cost-effectiveness acceptability curves. In Figure 2-A the points are spread between an incremental effect on the x-axis of -0.15 and +0.20, while on Figure 2-C all the points show an incremental effect on the x-axis above 0. The north-east quadrant is where the intervention is more costly but more effective and the north-west quadrant is where the intervention is more costly but less effective. Figure 2-B shows that the probability of the intervention to be cost-effective is 49% for a threshold of ∼€20000 (Portugal per-capita GDP) and Figure 2-D shows that the probability of the intervention be cost-effective is more than 70% using threshold of €1350 and 100% from a threshold of €2400. Table summarizes the sensitivity analysis based on different cost scenarios, the intervention remains cost-effective in all cost scenarios.

**Figure 2.**
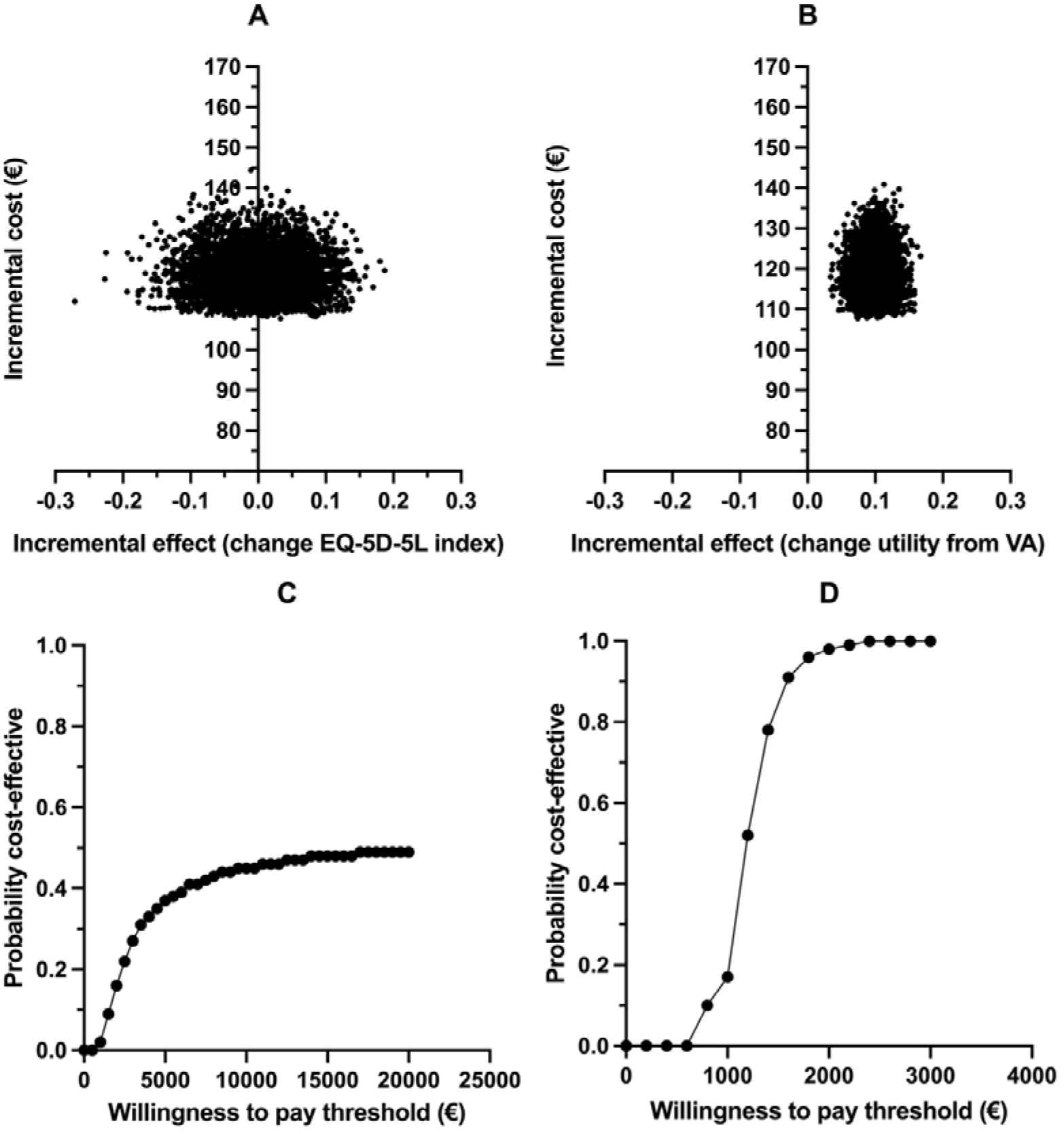
**Top row**: cost-effectiveness planes, A – based on EQ-5D-5L and B – based on near VA. **Bottom-row**, cost-effectiveness acceptability curves, C – based on EQ-5D-5L and D – based on near VA, generated from cost and effectiveness data.

## Discussion

In the current study we investigated the effect of a basic vision rehabilitation intervention and its cost-effectiveness. The study took place in a public hospital and was focused on a single interaction with an optometrist providing updated refractive correction, reading aids, basic training with the aid and instructions to reduce the effect of low vision in activities of daily living. The ability to perform activities that rely on vision - visual ability – was the main outcome measure and was assessed with the AI. As hypothesized, a basic vision rehabilitation intervention resulted in a significant improvement in visual ability. These results are in line with previous studies conducted in other countries [4, 18, 21, 26, 31, 55, 61-63]. In addition, there was an improvement in health-related quality-of-life as measured by the EQ-5D-5L, which is also in line with other studies [26, 62, 63]. The current basic vision rehabilitation intervention was cost-effective which is in line with studies investigation the cost-effectiveness of other type of rehabilitation interventions for people with VI [55, 56, 64-66].

### Clinical impact of rehabilitation

The improvement seen in our participants’ visual ability suggests that they experienced less difficulties to perform activities of daily living, which provides evidence of the benefits of vision rehabilitation for people with visual impairment [4, 5, 61]. The difference between the current study and previous studies is that others tended to included comprehensive rehabilitation with different professionals (e.g., occupational therapist, social worker), and multiple interactions with the patients to improve aspects such as mobility or at home training [4, 5, 61, 67, 68]. Therefore, the current study gives fresh evidence that a simple but structured intervention produces measurable improvements in everyday functioning for people with VI.

Improvements in visual ability can be explained by an overall improvement in vision function and, eventually, an optimization of the remaining vision. The improvement in acuity at distance (overall approximately 2 letters) can be considered modest but reasonable if we consider that with some treatments such as anti-VEGF injections the improvement in distance VA is in the range 1-5 letters [69, 70]. Most of our participants were unable to read common prints sizes, such as personal mail, before rehabilitation. The intervention caused significant improvements in near VA and critical print size which reduced the reading difficulties reported in the AI. This was an expected finding because vision aids should improve near vision tasks and in particularly reading [18, 21, 26, 71-73]. In short, changes in visual ability detected after the basic vision rehabilitation intervention can be explained by improvements in near vision tasks achieved with the correct use of the prescribed magnification.

### Cost-effectiveness- discussion of the economical evaluation

The cost of the basic vision rehabilitation intervention was lower than the costs reported by other studies [32]. These results can be explained by the normal differences in costs of healthcare workforce and products in different countries. Bray et al. has found that interventions for near vision activities tend to be cost-effective independently of the type of magnifiers [55]. Our findings are in line with these results.

ICERs calculations for different utilities (Table) and costs scenarios (Table), show that our intervention was always cost-effective assuming a threshold equal to the Portuguese per-capita GDP. However, when using the EQ-5D index we found that the probability of the intervention to be cost-effectiveness was roughly 49% - which indicates uncertainty around these estimates (Figure 2: B). Based on the near vision ophthalmic utility value the intervention is more cost-effective than with EQ-5D-5L index value and there was less uncertainty around the estimates (Figure 2-D). This may be related with the fact that the near acuity based ophthalmic utility value captures the actual near vision improvements. The EQ-5D-5L index has been considered to have low responsiveness to the effects of vision rehabilitation and, therefore, our results for the cost-effectiveness have limitations [38, 74]. That is, the fact that we used the EQ-5D-5L, an instrument with limited responsiveness to vision rehabilitation is a limitation to the results of the current study.

In ophthalmologic interventions a literature review concluded that, by conventional standards, the majority of interventions are cost-effective and found a median threshold value of $5,219/QALY (∼ €4571.24/QALY), range from $746/QALY to $6.5 million/QALY [75]. Recent findings in vision rehabilitation services in England found that for values between £13,000 (∼€15,423) and £30,000 (∼€35,591) per QALY, in-house VR has a high probability of being cost-effective under a social care perspective. Although, the probability of being cost-effective was lower when a healthcare perspective was used [64]. Assuming a healthcare perspective, which is expected to capture only part of the benefits of vision rehabilitation, the basic intervention performed as part of the current study can be considered cost-effective as shown by the different costs and utility scenarios investigated.

We consider that the COVID-19 pandemic was a significant barrier for recruiting and for retaining participants in this study which led to its first limitation – the *n* was smaller than expected and can be considered small. The small sample affected particularly the cost-effectiveness analysis where results for QALY gains and losses for the groups were very “noisy”-this limits the strength of the findings. Another limitation was the fact that the research person collecting the data (author LHM) was not “blinded” for the allocation of the participants. That might have caused bias during data collection; although, the researcher was always aware of this fact and did all possible to control any bias. These limitations should be addressed in future work in the field.

## Conclusion

Results of the current study show that a basic vision rehabilitation intervention was clinically impactful and cost-effective. A single interaction patient-optometrist led to immediate meaningful improvements for the patient that were retained over time. These findings are important to clinical and rehabilitation practices and for planning vision rehabilitation services.

## Data Availability

All data produced in the present study are available upon reasonable request to the authors

## a) Funding/Support

Ophthalmic lenses and part of the magnifiers were supported by Essilor Portugal and FCT Strategic Funding UID/FIS/04650/2013. AFM was funded by the faculty of Health and Life Sciences at Linnaeus University.

## b) Disclosures

The authors report no conflicts of interest and have no proprietary interest in any of the materials mentioned in this article.

## c) Other Acknowledgments

The current study is part of a clinical trial which has started in March 2017 (registration number: ISRCTN10894889 (https://doi.org/10.1186/ISRCTN108948899). We acknowledge the hospital where the study took place for the availability of the space. We also would like to acknowledge staff from the department of ophthalmology, including all ophthalmologists, for their collaboration in the recruitment process. We also acknowledge Dr. João Linhares, Professor Rui Santana, Dr Pedro Lima Ramos for their support during this project.

## Notes

### Competing Interest Statement

The authors have declared no competing interest.

### Clinical Trial

ISRCTN10894889

### Clinical Protocols

https://doi.org/10.1186/ISRCTN108948899

### Author Declarations

The study was approved by the Ethics Committee for Life Sciences and Health of the University of Minho (SECVS 147/2016), and by the Hospital Santa Maria Maior's ethics committee. Portuguese data protection authority approval number: 7012/ 2017.

## References

1. WHO, World Report on Vision. World Health Organization, 2019.

2. Burton, M.J., et al., The Lancet Global Health Commission on Global Eye Health: vision beyond 2020. Lancet Glob Health, 2021. 9(4): p. e489–e551.

3. Devlin, N.J., et al., Valuing health-related quality of life: An EQ-5D-5L value set for England. Health Econ, 2018. 27(1): p. 7–22.

4. Pearce, E., M.D. Crossland, and G.S. Rubin, The efficacy of low vision device training in a hospital-based low vision clinic. Br J Ophthalmol, 2011. 95(1): p. 105–8.

5. Stelmack, J.A., et al., Outcomes of the Veterans Affairs Low Vision Intervention Trial II (LOVIT II): A Randomized Clinical Trial. JAMA Ophthalmol, 2017. 135(2): p. 96–104.

6. Rees, G., et al., A randomised controlled trial of a self-management programme for low vision implemented in low vision rehabilitation services. Patient Educ Couns, 2015. 98(2): p. 174–81.

7. Marques, A.P., et al., The use of informal care by people with vision impairment. PLoS ONE 2018. 13(6): p. e0198631.

8. Marques, A.P., et al., Productivity Losses and Their Explanatory Factors Amongst People with Impaired Vision. Ophthalmic Epidemiol, 2019: p. 1–15.

9. Hernandez-Moreno, L., et al., Is perceived social support more important than visual acuity for clinical depression and anxiety in patients with age-related macular degeneration and diabetic retinopathy? Clin Rehabil, 2021: p. 269215521997991.

10. Rabiee, P., et al., Community-based vision rehabilitation provision in England. British Journal of Visual Impairment, 2016. 34(3): p. 248–261.

11. Taylor, H.R., M.L. Pezzullo, and J.E. Keeffe, The economic impact and cost of visual impairment in Australia. Br J Ophthalmol, 2006. 90(3): p. 272–5.

12. Cruess, A.F., et al., The cost of vision loss in Canada. 2. Results. Can J Ophthalmol, 2011. 46(4): p. 315–8.

13. Wittenborn, J.S., et al., The economic burden of vision loss and eye disorders among the United States population younger than 40 years. Ophthalmology, 2013. 120(9): p. 1728–35.

14. Marques, A.P., et al., Global economic productivity losses from vision impairment and blindness. EClinicalMedicine, 2021. 35: p. 100852.

15. Coyne, K.S., et al., The impact of diabetic retinopathy: perspectives from patient focus groups. Fam Pract, 2004. 21(4): p. 447–53.

16. Langelaan, M., et al., Impact of visual impairment on quality of life: A comparison with quality of life in the general population and with other chronic conditions. Ophthalmic Epidemiol, 2007. 14(3): p. 119–26.

17. Brown, J.C., et al., Characterizing functional complaints in patients seeking outpatient low-vision services in the United States. Ophthalmology, 2014. 121(8).

18. Stelmack, J.A., et al., Outcomes of the Veterans Affairs Low Vision Intervention Trial (LOVIT). Arch Ophthalmol, 2008. 126(5): p. 608–17.

19. WHO, International Classification of Functioning, Disability and Health, 2001. Geneva. World Health Organization, 2001.

20. Massof, R.W., et al., The activity inventory: an adaptive visual function questionnaire. Optom Vis Sci, 2007. 84(8): p. 763–74.

21. Stelmack, J.A., et al., The effectiveness of low-vision rehabilitation in 2 cohorts derived from the veterans affairs Low-Vision Intervention Trial. Arch Ophthalmol, 2012. 130(9): p. 1162–8.

22. Stronks, H.C., et al., Visual task performance in the blind with the BrainPort V100 Vision Aid. Expert Rev Med Devices, 2016. 13(10): p. 919–931.

23. Crossland, M.D., R.S. Silva, and A.F. Macedo, Smartphone, tablet computer and e-reader use by people with vision impairment. Ophthalmic Physiol Opt, 2014. 34(5): p. 552–7.

24. Smith, T.M., I. Hong, and T.A. Reistetter, Responsiveness of the Revised Low Vision Independence Measure (LVIM-R). Am J Occup Ther, 2020. 74(5): p. 7405205040p1–7405205040p11.

25. Malkin, A.G., J.E. Goldstein, and R.W. Massof, Multivariable Regression Model of the EuroQol 5-Dimension Questionnaire in Patients Seeking Outpatient Low Vision Rehabilitation. Ophthalmic Epidemiol, 2017. 24(3): p. 174–180.

26. Binns, A.M., et al., How effective is low vision service provision? A systematic review. Surv Ophthalmol, 2012. 57(1): p. 34–65.

27. Hernández-Moreno, L., et al., Visual and psychological outcomes in patients with and without low vision diagnosed with similar eye diseases - initial results. Invest Ophth Vis Sci, 2018. 59(9): p. 3411–3411.

28. Senra, H., et al., Psychological and Psychosocial Interventions for Depression and Anxiety in Patients with Age-Related Macular Degeneration–A Systematic Review. Am J Geriatr Psychiatry, 2019. 27(8): p. 755–773.

29. Ramos, P.L., et al., Predicting participation of people with impaired vision in epidemiological studies. BMC Ophthalmol, 2018. 18(1): p. 236.

30. Ramos, P.L., et al., Prevalence and causes of vision impairment in Norwest Portugal: a capture and recapture study. 2021, medRxiv.

31. Stelmack, J.A., et al., VA LOVIT II: a protocol to compare low vision rehabilitation and basic low vision. Ophthalmic Physiol Opt, 2012. 32(6): p. 461–71.

32. Stroupe, K.T., et al., Economic evaluation of low-vision rehabilitation for veterans with macular diseases in the US department of veterans affairs. Jama Ophthalmol, 2018. 136(5): p. 524–531.

33. Lam, N. and S.J. Leat, Barriers to accessing low-vision care: the patient’s perspective. Can J Ophthalm, 2013. 48(6): p. 458–462.

34. O’Connor, P.M., L.C. Mu, and J.E. Keeffe, Access and utilization of a new low-vision rehabilitation service. Clin Exp Ophthalmol, 2008. 36(6): p. 547–52.

35. Crossland, M.D. and J.H. Silver, Thirty years in an urban low vision clinic: changes in prescribing habits of low vision practitioners. Optom Vis Sci, 2005. 82(7): p. 617–22.

36. Virgili, G., et al., Reading aids for adults with low vision. Cochrane Db Syst Rev, 2013(10): p. Cd003303.

37. Virgili, G., et al., Reading aids for adults with low vision. Cochrane Db Syst Rev, 2018(4).

38. Hernandez-Moreno, L., et al., Cost-effectiveness of basic vision rehabilitation (The basic VRS-effect study): study protocol for a randomised controlled trial. Ophthalmic Physiol Opt, 2020.

39. Guerreiro, M., et al., Adaptação à população portuguesa da tradução do Mini Mental State Examination (MMSE). Revista Portuguesa de Neurologia, 1994. 1(9): p. 9–10.

40. Hernandez-Moreno, L., et al., The Portuguese version of the activity inventory. Invest Ophth Vis Sci, 2015. 56(7).

41. Macedo, A.F., et al., Visual and health outcomes, measured with the activity inventory and the EQ-5D, in visual impairment. Acta Ophthalmol, 2017. 95(8): p. e783–e791.

42. Massof, R.W., et al., Visual disability variables. I: the importance and difficulty of activity goals for a sample of low-vision patients. Arch Phys Med Rehabil, 2005. 86(5): p. 946–53.

43. Massof, R.W., et al., Visual disability variables. II: The difficulty of tasks for a sample of low-vision patients. Arch Phys Med Rehabil, 2005. 86(5): p. 954–67.

44. Ferris, F.L., et al., New Visual-acuity charts for clinical research. Am J Ophthalmol, 1982. 94(1): p. 91–6.

45. Legge, G.E., et al., Psychophysics of reading. VIII. The Minnesota Low-Vision Reading Test. Optom Vis Sci, 1989. 66(12): p. 843–53.

46. Castro, C.T., C.S. Kallie, and S.R. Salomao, Development and validation of the MNREAD reading acuity chart in Portuguese. Arq Bras Oftalmol, 2005. 68(6): p. 777–83.

47. Baskaran, K., et al., Scoring reading parameters: An inter-rater reliability study using the MNREAD chart. PloS one, 2019. 14(6): p. e0216775–e0216775.

48. Herdman, M., et al., Development and preliminary testing of the new five-level version of EQ-5D (EQ-5D-5L). Qual Life Res, 2011. 20(10): p. 1727–36.

49. Oppe, M., et al., A Program of methodological research to arrive at the new international EQ-5D-5L valuation protocol. Value Health, 2014. 17(4): p. 445–53.

50. Ross, E.L., et al., Cost-effectiveness of Aflibercept, Bevacizumab, and Ranibizumab for Diabetic Macular Edema Treatment: Analysis From the Diabetic Retinopathy Clinical Research Network Comparative Effectiveness Trial. JAMA Ophthalmol, 2016. 134(8): p. 888–96.

51. Brown, G.C., Vision and quality-of-life. Transactions of the American Ophthalmological Society, 1999. 97: p. 473.

52. Brown, M.M., et al., Health care economic analyses and value-based medicine. Surv Ophthalmol, 2003. 48(2): p. 204–23.

53. INE. Produto interno bruto por habitante a preços correntes por localização geográfica. 2020 14 December 2021 17 February 2022]; Available from: https://www.ine.pt/xportal/xmain?xpid=INE&xpgid=ine_indicadores&contecto=pi&indOcorrCod=0009975&selTab=tab0.

54. Garber, A.M. and C.E. Phelps, Economic foundations of cost-effectiveness analysis. Journal of Health Economics, 1997. 16(1): p. 1–31.

55. Bray, N., et al., Portable electronic vision enhancement systems in comparison with optical magnifiers for near vision activities: an economic evaluation alongside a randomized crossover trial. Acta Ophthalmol 2017. 95(5): p. e415–e423.

56. Patty, N.J.S., M. Koopmanschap, and K. Holtzer-Goor, A cost-effectiveness study of ICT training among the visually impaired in the Netherlands. BMC Ophthalmol, 2018. 18(1): p. 98.

57. Bang, H. and H. Zhao, Median-Based Incremental Cost-Effectiveness Ratio (ICER). J Stat Theory Pract, 2012. 6(3): p. 428–442.

58. Reed, S.D., Statistical considerations in economic evaluations: a guide for cardiologists. Eur Heart J, 2014. 35(25): p. 1652–6.

59. David, A., A rating formulation for ordered response categories. Psychometrika, 1978. 43(4): p. 561–573.

60. Wright, B.D. and J.M. Linacre, Observations are always ordinal - measurements, however, must be interval. Arch Phys Med Rehabil, 1989. 70(12): p. 857–860.

61. Goldstein, J.E., et al., Clinically Meaningful Rehabilitation Outcomes of Low Vision Patients Served by Outpatient Clinical Centers. JAMA Ophthalmol, 2015. 133(7): p. 762–9.

62. Do, A.T., et al., Effectiveness of low vision services in improving patient quality of life at Aravind Eye Hospital. Indian Journal of Ophthalmology, 2014. 62(12): p. 1125–1131.

63. Kaltenegger, K., et al., Effects of home reading training on reading and quality of life in AMD-a randomized and controlled study. Graefes Arch Clin Exp Ophthalmol, 2019. 257(7): p. 1499–1512.

64. Longo, F., et al., Cost-Effectiveness of In-House Versus Contracted-Out Vision Rehabilitation Services in England. Journal of Long-term Care, 2020: p. 118–130.

65. Taylor, J., et al., The p-EVES study design and methodology: a randomised controlled trial to compare portable electronic vision enhancement systems (p-EVES) to optical magnifiers for near vision activities in visual impairment. Ophthalmic Physiol Opt, 2014. 34(5): p. 558–72.

66. van der Aa, H.P.A., et al., Economic evaluation of stepped-care versus usual care for depression and anxiety in older adults with vision impairment: randomized controlled trial. BMC Psychiatry, 2017. 17(1): p. 280.

67. Lamoureux, E.L., et al., The effectiveness of low-vision rehabilitation on participation in daily living and quality of life. Invest Ophthalmol Vis Sci, 2007. 48(4): p. 1476–82.

68. Silva, M.B.R., et al., Quality of life of patients attending a low-vision rehabilitation service in Brazil. Invest Ophth Vis Sci, 2018. 59(9): p. 1066–1066.

69. Wykoff, C.C., et al., SAVE (Super-dose anti-VEGF) trial: 2.0 mg ranibizumab for recalcitrant neovascular age-related macular degeneration: 1-year results. Ophthalmic Surg Lasers Imaging Retina, 2013. 44(2): p. 121–6.

70. Wallsh, J.O. and R.P. Gallemore, Anti-VEGF-Resistant Retinal Diseases: A Review of the Latest Treatment Options. Cells, 2021. 10(5): p. 1049.

71. van Nispen, R.M., et al., Applying multilevel item response theory to vision-related quality of life in Dutch visually impaired elderly. Optom Vis Sci, 2007. 84(8): p. 710–20.

72. Margrain, T.H., Helping blind and partially sighted people to read: the effectiveness of low vision aids. Br J Ophthalmol, 2000. 84(8): p. 919–21.

73. Hinds, A., et al., Impact of an interdisciplinary low vision service on the quality of life of low vision patients. Br J Ophthalmol, 2003. 87(11): p. 1391–6.

74. Malkin, A.G., et al., Responsiveness of the EQ-5D to the effects of low vision rehabilitation. Optom Vis Sci, 2013. 90(8): p. 799–805.

75. Brown, G.C., et al., Value-based medicine and ophthalmology: an appraisal of cost-utility analyses. Trans Am Ophthalmol Soc, 2004. 102: p. 177-85; discussion 185-8.

